# Laparoscopic Assisted Vaginal Hysterectomy Verses Abdominal Hysterectomy: A systematic review and metanalysis

**DOI:** 10.1101/2022.08.21.22279030

**Authors:** Esraa Menshawey, Rahma Menshawey

## Abstract

**OBJECTIVE:** To examine studies that explored the differences between laparoscopic assisted vaginal hysterectomy (LAVH) and total abdominal hysterectomy (TAH) in endometrial cancer (EC) patients, and to determine which surgical intervention has better outcomes.

**DATA SOURCES:** Electronic search of the following databases was performed; Google Scholar, PubMed/Medline, Wiley, Web of Science, Cochrane Library, Embase, and EBSCO Host.

**METHODS of STUDY SELECTION:** All full English articles in the form of randomized controlled trials (RCT), prospective cohort (PC), and retrospective cohort (RC) comparing LAVH and TAH outcomes in endometrial cancer patients was included in this study. A complete search of the literature comparing the outcomes of LAVH and AH in EC patients. This study was registered in PROSPERO [ID: CRD42021225509] and follows PRISMA and MOOSE guidelines. Outcomes included length of hospital stay, surgical duration, complications, blood transfusion requirements, and blood loss.

**TABULATION:** ROBINS-1, ROB 2.0, and ROBVIS was used to assess the risk of bias. Statistical tests used included relative risk (RR) for dichotomous and standard mean difference (SMD) for continuous variable. A *P* value less than 0.05 was considered significant. A forest plot was used to visually demonstrate the analyses for all outcomes.

**INTEGRATION and RESULTS:** A total of 13 articles (total cohort n=14,803) were included in the systematic review and metanalysis. The total cohort for LAVH patients was n=1845 and n=12,958 for TAH. Patients who underwent a TAH had significantly higher risk of complications [RR = 0.547, *p<*0.001], greater risk for blood transfusion [RR = 0.349, *p<*0.033], more blood loss [SMD = −3.256, *p<*0.001], and longer hospital stay [SMD = −1.351, *p<*0.001]. LAVH patients had longer operating time [SMD= 1.103, *p<*0.001] compared TAH patients.

**CONCLUSION:** LAVH presented with lower of hospital stay, complications, amount of blood loss, and blood transfusion requirements when compared to TAH. LAVH in the appropriate setting and skills may be a safer alternative than TAH.

## Introduction

Uterine cancer is the sixth most common female malignancy in the world [1]. More than 90% of all uterine cancers involve the endometrium. The first hysterectomy (laparotomy) was performed by Charles Clay back in 1843, England [2]. In 1988, Harry Reich performed the first laparoscopic hysterectomy [2]. Till now, the laparotomy approach to a hysterectomy tends to predominate approach despite technological advancements. Each surgical technique has its downsides and benefits, which is why comparing of surgical approaches is needed to find the safest method with the best results and outcomes. In this systematic review and meta-analysis, we examined other literature which studied the differences between laparoscopic assisted vaginal hysterectomy (LAVH) and total abdominal hysterectomy (TAH) in females with endometrial carcinoma.

## Methods

This systematic review and meta-analysis is registered by PROSPERO [ID: CRD42021225509].

### A. Search strategy

We thoroughly performed an electronic search from November 2020 till July 2021 using the following search engines; Google Scholar, PubMed/Medline, Wiley, Web of Science, Cochrane Library, Embase, and EBSCO Host. Key words that were used in the search included Laparoscopic assisted vaginal hysterectomy, total abdominal hysterectomy, and endometrial carcinoma. We searched articles from the beginning of 1990 till early 2021. Articles were included based on the inclusion criteria. Articles were scanned based on title and abstract; any duplicates were removed. In case of missing data, authors of the articles were contacted to provide further details when necessary. This systematic review and metanalysis was conducted in accordance with PRISMA guidelines (preferred reporting items for systematic reviews and meta-analyses).

### B. Data collection and analysis

Two authors (E.M and R.M) screened the data and independently assessed the risk of bias.

### C. Inclusion Criteria

I. Intervention – only articles which compared laparoscopic assisted vaginal hysterectomy and total abdominal hysterectomy were included.
II. Study type – All full English articles in the form of randomized controlled trials (RCT), prospective cohort (PC), and retrospective cohort (RC) were included in our study.
III. Type of participants – adult females requiring a hysterectomy for any stage of endometrial carcinoma.

### D. Exclusion Criteria

All non-English articles, and grey articles or conference papers were not included. Any study that compared other forms of hysterectomy (ex. Robotic assisted, etc.) for female patients with cancer other than endometrial carcinoma were not included. Any study that had not included the outcomes assessed in this study was also not included.

### E. Outcomes

Length of hospital stay, complications (pre-operative, intra-operative, and post-operative), quantity of blood loss (mL), the need for blood transfusion, and total duration for surgical intervention were all the outcomes analysed. The rate of recurrence and conversion to laparotomy approach was an outcome we documented but did not include it in the metanalysis due to lack of substantial of data. (Table 1)

### F. Assessment and Risk of Bias

All eligible articles included in this study were used for both qualitative and quantitative analysis. Tools used for bias screening followed the guidelines of the *Cochrane Handbook for Systematic Reviews of Interventions* chapter 8. [3] For all non-randomized control trial studies (prospective and retrospective cohort), risk of bias was assessed using the ROBINS-1 (Cochrane method for systematic reviews) tool, while the randomized controlled trials were evaluated for risk of bias using the ROB 2.0 tool. Visual demonstration of the risk of bias for all study types was depicted using the ROBVIS tool (Cochrane method for systematic reviews).

### G. Statistical analysis and tools

As mentioned earlier, tools for risk of bias assessment included ROBINS-1, ROB 2.0, and ROBVIS. Data collection for general information (author, year, intervention, number of cohorts, etc.) was imputed into a Microsoft Excel Sheet. For statistical analyses, MedCalc Statistical Software Version 19.2.6 (MedCalc software, Ltd, Ostend, Belgium) was used. Regarding data input and analysis, relative risk (RR) was used for any dichotomous variables (Complications and need for blood transfusion) and standard mean difference (SMD) was used for continuous variables (hospital stay, duration of surgery, and amount of blood loss). Effects estimates were reported with 95% confidence interval. A *P* value less than 0.05 was considered significant. A forest plot was used to visually demonstrate the analyses for all outcomes.

## Results

### A. Patients and Intervention

Altogether, there were 14,803 patients who underwent a hysterectomy, 1845 of which had a LAVH and the other 12,958 had underwent a TAH. (**Table 1**)

### B. Electronic search

When searching the keywords, a total of 32,889 articles were found altogether in the sources mentioned, which underwent initial screening. After removal of duplicates, screening of titles, and assessing the eligibility of the articles based on the inclusion criteria, a total of 13 articles were used for both quantitative and qualitative analyses (PRISMA diagram – **Figure 1**). Of the 13 articles, four were retrospective cohort (Tollund, Palomba, Leiserowitz, and Frigerio), three were prospective cohort (Devaja, Howard, and Kalogiannidis), and six were randomized controlled trials (Falcone, Malur, Fram, Malzoni, Tehranian, and Atabekoglu).

**Figure 1:**
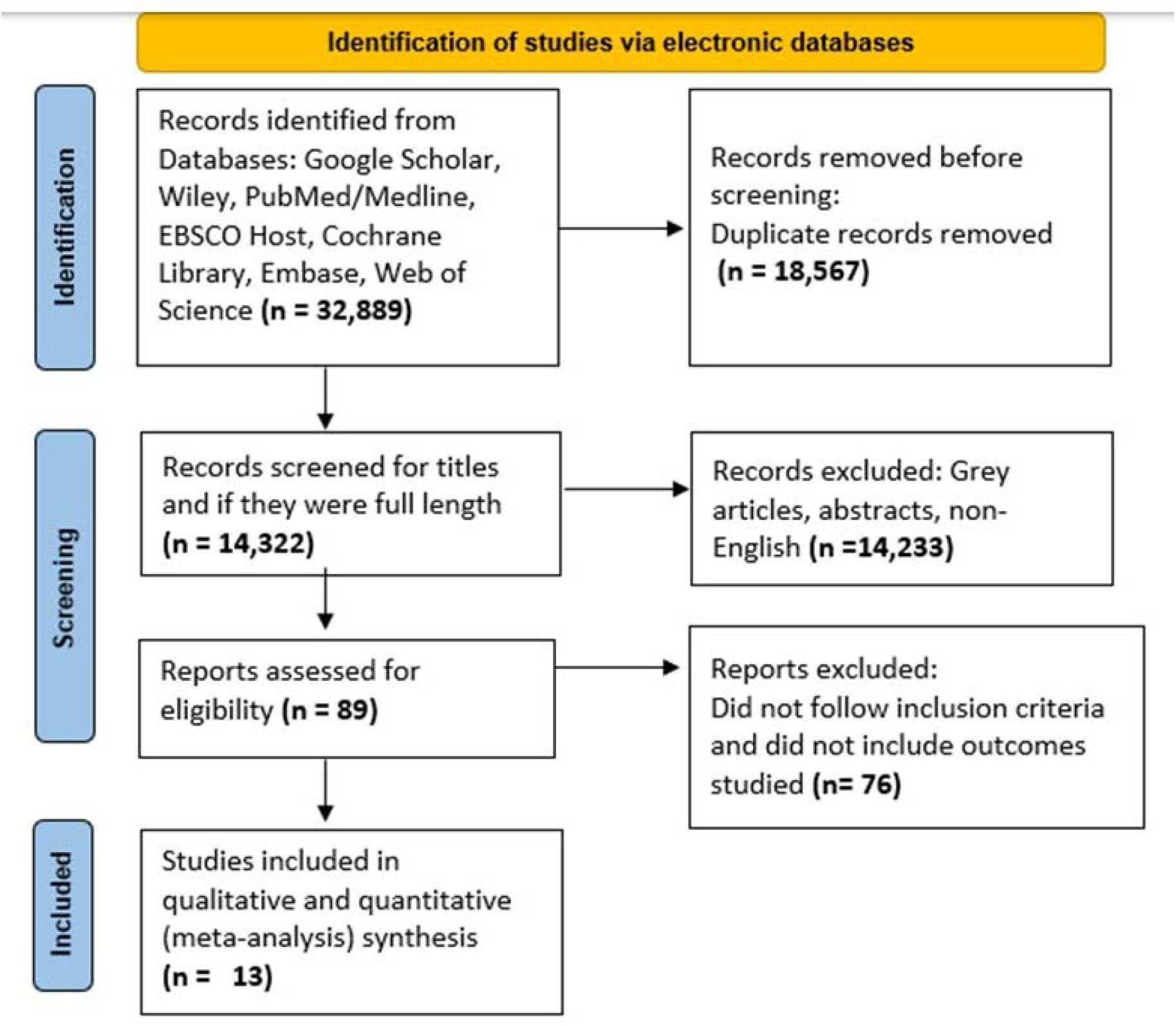
PRISMA Flowchart

### C. Risk of Bias

Of the six randomized control trials (**Figure 2**), three had a low risk of bias (Fram, Malzoni, and Tehranian), two (Atabekoglu and Falcone) had an overall high risk of bias due to deviation from the intended intervention, and one (Malur) had a moderate degree of bias for the same reason as the high risk of bias articles. Deviation from the intended intervention was considered when the patient was initially randomly selected for one intervention, but due to patient related factors had to be switched to the other intervention either pre-operatively or intra-operatively.

**Figure 2:**
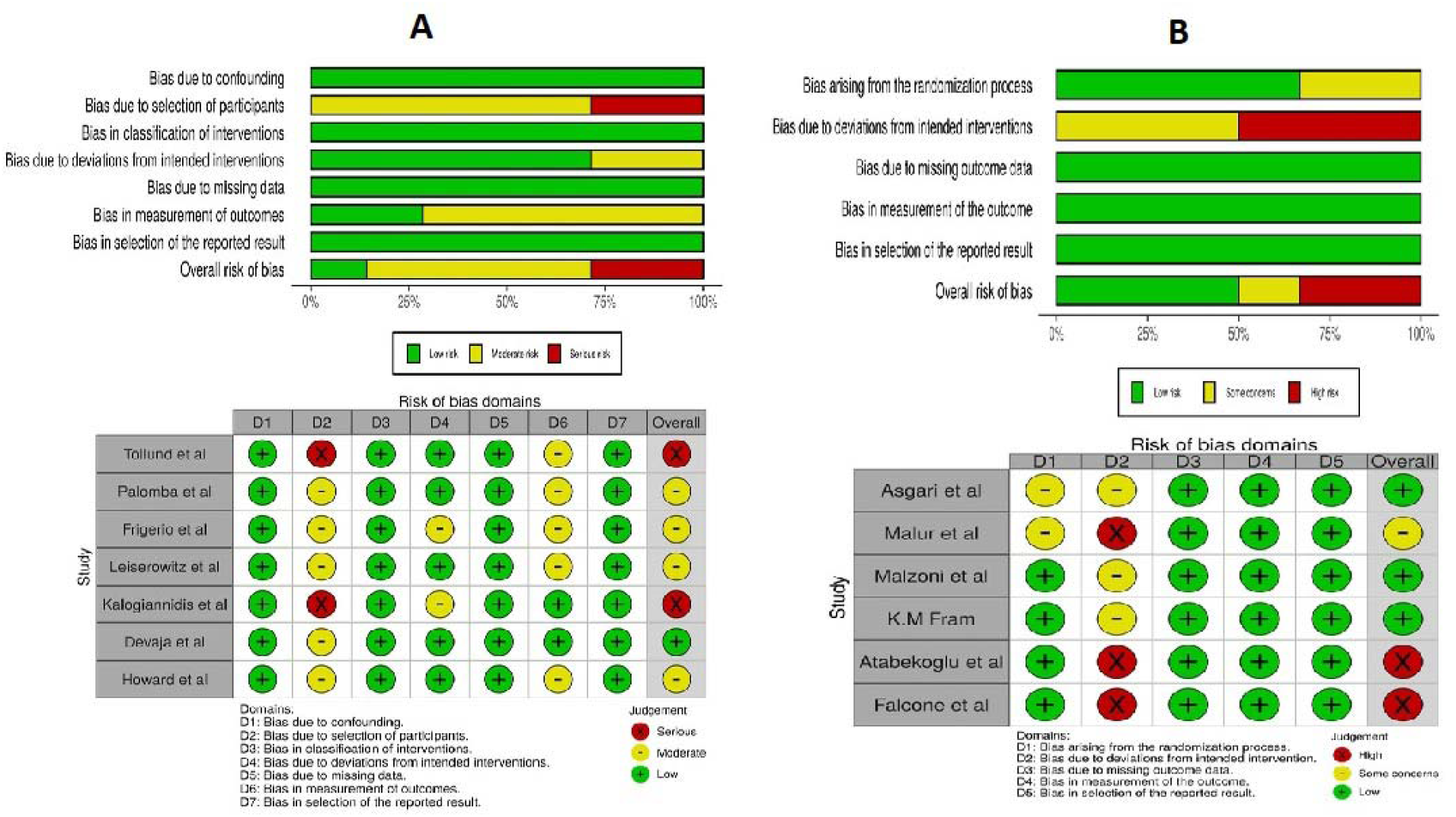
Visual demonstration of risk of bias assessment using the ROBVIS tool. (A) is the risk of bias assessment for both prospective and retrospective cohort. (B) is the risk of bias assessment for randomized control trials.

Of the seven prospective and retrospective cohorts (**Figure 2**), two studies (Tollund and Kalogiannidis) were considered highly biased in terms of patient selection, four were moderately bias for the same reason, and one study (Devaja) was not considered bias. This may be because retrospective and similarly prospective cohorts are commonly more prone to selection bias due to the available selection of already existing patient data, which gives the researcher the ability to pick what cases they presume as satisfactory for the study without any form of randomization. [4]

### D. Outcome results

#### D.1 Dichotomous Variables: (**Figure 3**)

I. Complications: Any complication, whether pre-operative, intra-operative, or post-operative was considered in the analysis. The relative risk for the complication was 0.547 (CI =0.466-0.642) and a *P value < 0*.*001* (significant). Test for heterogeneity was P=0.1790. This indicates that TAH had greater complications than LAVH.

**Figure 3:**
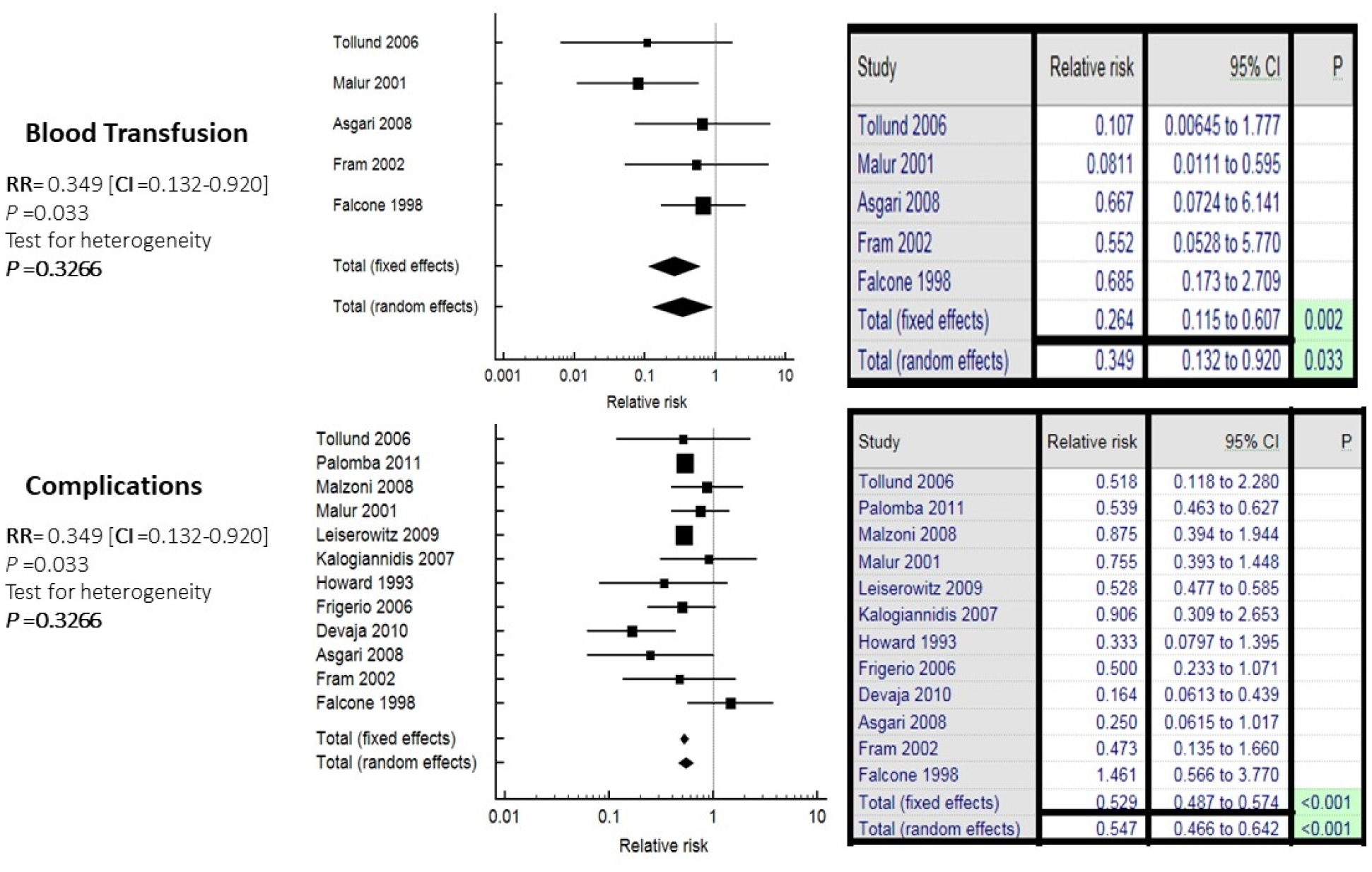
Forest plot representation for dichotomous variables (complications and need for blood transfusion).
II. The need for Blood Transfusion: The relative risk for patients who needed a blood transfusion was 0.349 (CI = 0.132-0.920), with a *P value of 0*.*033* (significant) and the test of heterogeneity was P=0.03266. This means that patients who underwent a TAH were 34.9% at a greater risk of requiring blood transfusion, or in other words, those who had a LAVH were 65.1% less likely to need a blood transfusion.

#### D.2 Continuous variables: (**Figure 4**)

I. Length of Hospital Stay: Hospital stay SMD = −1.351 (CI= −2.046 to −0.657) with a *P value <0*.*001* (significant). This means that patients who underwent a TAH had longer hospital stay than those who underwent a LAVH.

**Figure 4:**
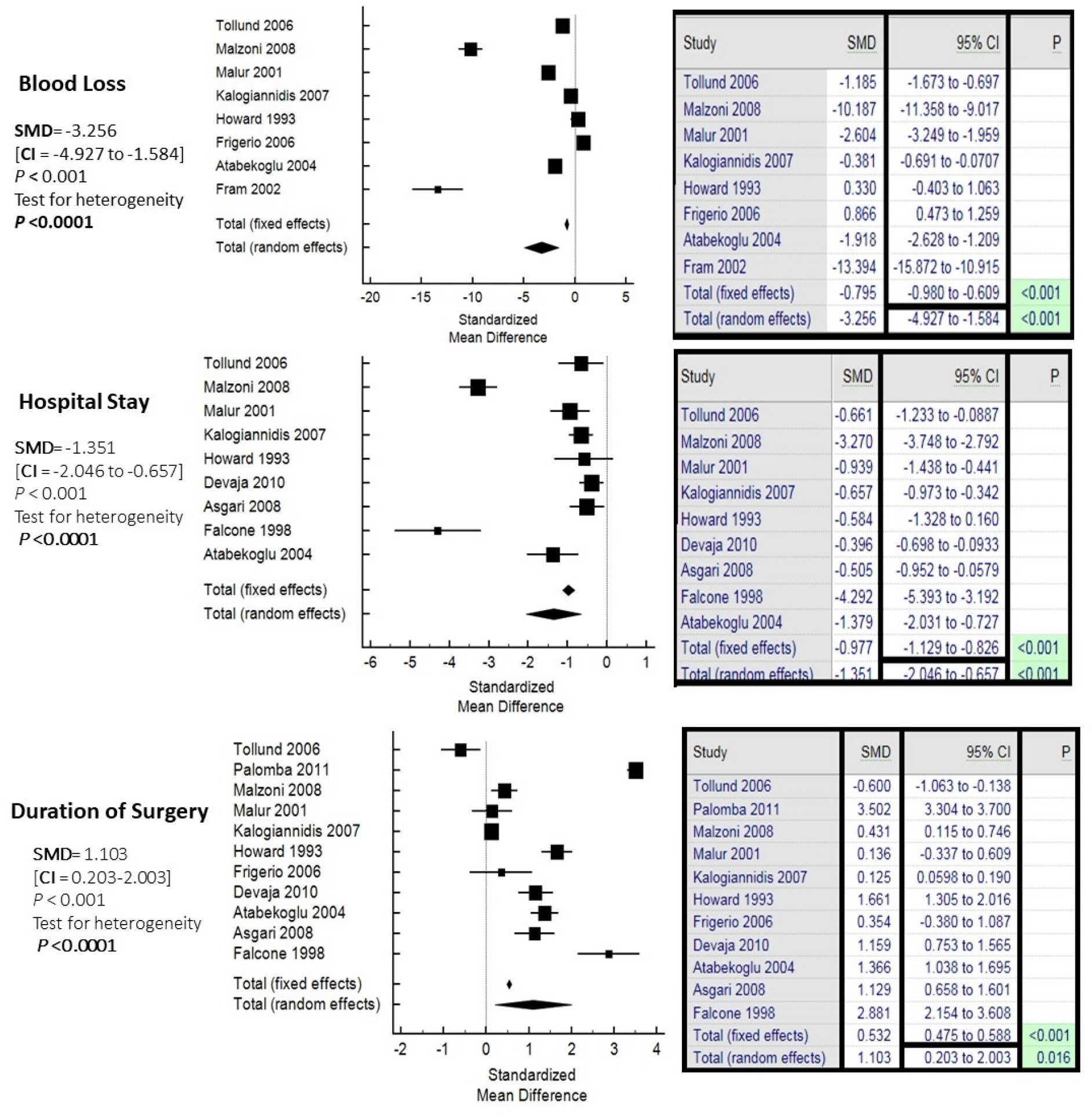
Forest plot representation for continuous variable (hospital stay, duration of surgery, and blood loss).
II. Total Blood Loss: Blood loss SMD = −3.256 (CI= −4.927 to −1.584) with a *P value <0*.*001* (significant). Therefore, TAH patients had greater blood loss than those who underwent a LAVH.
III. Duration of Surgery: Total time to perform surgery SMD =1.103 (CI= 0.203-2.003) with a *P value <0*.*001* (significant). Therefore, patients who underwent LAVH had longer operating time than those who had a TAH.

Overall, patients who underwent a LAVH had a shorter hospital stay and longer operating time. Those who underwent a TAH were more likely to develop complications, had a greater risk of requiring blood transfusion, and lost more blood.

## Discussion

A hysterectomy may either be an elective or mandatory procedure. In cases of endometrial carcinoma, it is generally preferred to perform a hysterectomy to prevent further progression and complications of malignancy. In this systematic review, we specifically looked at studies comparing LAVH and TAH in EC patients. A total of 13 articles were included in the metanalysis.

TAH is the classical method for a hysterectomy, despite all the novel hysterectomy techniques. Leiserowitz et al pointed out that between the year 1997 and 2001, only 8% of EC patients preferred to have a LAVH, which was associated with a younger age group and patients who had private insurance [5]. Other studies have attributed the higher rates of TAH to hospital resources and exceptional skills that are needed for LAVH [6–8] [9]. Overall, this study analyzed the results of 1,845 LAVH patients and 12,958 TAH.

Surgical complications can either present pre/intra/ or post-operatively. In our metanalyses, TAH patients showed a significantly higher risk of developing complications than LAVH patients [RR= 0.547, p<0.001]. Fram et al documented less intra- and post-operative complications in the laparoscopic group [10]. The major complications that were reported included urinary tract infections (UTI), pulmonary embolism (PE), anemia, fever, tachyarrhythmias, and wound infection [11, 12]. Kalogiannidis et al reported only 7% of all laparoscopic patients had post-operative complications [13], meanwhile Falcone et al pointed out that, in Sweden, 95% of hysterectomies are preformed via a laparotomy (open abdominal) approach. [14]

Abdominal Laparotomy approaches are typically associated with longer hospital stays compared to laparoscopic approaches as demonstrated by our study [p<0.001]. Kalogiannidis et al documented a median hospital duration of five days for LAVH and eight days for TAH [13]. Similarly, Howard et al noted a difference in hospital stay duration by two whole days (TAH with the longer stay) [15]. It should be noted that, the length of hospital stay may vary among different hospitals and different countries where the procedure took place. For example, Kalogiannidis et al noticed a difference in hospital stays between American and European hospitals, which was attributed to hospital guidelines and health insurance status [13]. In terms of the duration of surgery, the laparoscopic approach took a significantly longer amount of time than the TAH [SMD=1.103, p<0.001]. This is in accordance with Devaja et al, who documented that LAVH took almost one hour longer than the TAH [12]. Palomba et al also noted similar results [16]. The laparoscopic approach tends to involve more steps and requires more equipment in the operating theatre, thereby extending the duration till its completed. It should be noted that operating time may be affected by the surgical skills of the surgeon and the complexity of the case.

The results for the total amount of blood lost and the need for blood transfusion are directly related, as the need for blood transfusion increases with the larger amount of blood lost during the operation. TAH uses wider incisions and involves the cutting of highly vascularized structures, which may explain the greater amount of blood loss during the operation, and subsequently increase the risk of needing a transfusion, as demonstrated in this study. Some studies documented blood loss and transfusion as a surgical complication and concluded that laparoscopic had much less complications overall [5]. However, the study by Howard et al reports higher blood loss in the laparoscopic approach and stated that it was the result of ligating the uterine vessels vaginally [15]. Similarly, Frigerio et al noticed greater blood loss in the LAVH group, which may be attributed to the technique and the skill level of whomever performed the surgery [17]. Overall, most of the studies did present with greater blood loss and higher risk of blood transfusion for the TAH group.

There were a few outcomes studied by some of the selected articles which were not included in the metanalysis that should be noted. This included the conversion from laparoscopic to a laparotomy approach and the recurrence of malignancy. There were two studies that demonstrated significant conversion rate to laparotomy. Palomba et al attributed this rate of conversion to the stage of EC, while Leiserowitz et al stated that the conversion was the result of significant perioperative complications [5, 16]. As for the recurrence rate of malignancy, there was no significant difference between the two groups, but studies mentioned that the recurrence of malignancy (local or distant) may be related to the stage and quality of the technique [5, 9, 16].

## Conclusion

Endometrial carcinoma (EC) is one of the most common female malignancies worldwide. Hysterectomy is the mainstay treatment EC. There are numerous surgical approaches to performing a hysterectomy, each of which has its own benefits and risks. Overall, from this systematic review and metanalysis, it can be noted that, LAVH has greater benefits in terms of hospital stay, complications, amount of blood loss, and blood transfusion requirements compared to TAH. Throughout the literature, TAH tends to be the most preformed surgery compared to LAVH due to the level of expertise and skills required to perform the latter surgery as well as the resources required. The evidence supports the advantages of LAVH as a better alternative compared to TAH. When given the appropriate setting, skill level, and resources, LAVH can become a more suitable surgical approach for patients with endometrial carcinoma. Future studies with larger LAVH cohorts should be conducted. The results of this study could be used to update or develop guidelines and protocols.

## Supporting information

file:///C:/Users/Esraa/Desktop/Final%20table%20for%20systematic%20review.pdf

## Data Availability

All data produced in the present work are contained in the manuscript.

## Limitations

The present meta-analysis has a few limitations that warrant consideration when analyzing the results. The outcomes analyzed in this systematic review are focused on a limited number of variables (complications, blood loss, hospital stay, need for blood transfusion, and duration of intervention). The selection of variables was dependent on how common the parameters were reported. Including other outcomes such as cost effectiveness, quality of life post-operatively, pain management, morbidity, and mortality may aid in enhancing the quality of the study. Another limitation would be the different sample sizes among the included literature. Furthermore, even with the basic guidelines on how to perform these operations, institutions might have managed the intervention and post-operative follow-up differently. Also, results of the meta-analyses may have been heavily weighted by the articles with very large cohort sizes.

### Ethical Considerations

This study does not contain any studies with human or animal participants performed by any of the authors.

### Funding

This systematic review and metanalysis did not receive any funding from either governmental or non-governmental institutions.

## Acknowledgements

None.

## Author Contributions

All the listed authors have met the authorship criteria recommended by ICJME standards. E.M conceived the idea. EM and RM performed the database search, data collection, extraction, and statistical analysis. Assessment for bias was done by each author individually. The final manuscript revision, writing, and proofreading was done by both authors.

## Figure and Table Legend

**Table 1:** General information and reported outcomes collected from articles selected for the meta-analysis.

