## Supplementary material for "Laparoscopic Assisted Vaginal Hysterectomy Verses Abdominal Hysterectomy: A systematic review and metanalysis": file:///C:/Users/Esraa/Desktop/Final%20table%20for%20systematic%20review.pdf

| **First Author** | **Year** | **Country** | **Cohort** | **LAVH** | **TAH** | **H.S (Days)** | **Surgery Time (mins.)** | **Complications** | **B.L (mL)** | **B. T** | **C.** | **Recurrence** |
| --- | --- | --- | --- | --- | --- | --- | --- | --- | --- | --- | --- | --- |
| Tollund | 2006 | Denmark | 86 | 28 | 58 | 2.7 (3)  5.4 (5) | 91 (58-135)  92 (54-255) | 2 vs 8 | 184 (50-1200)  379 (50-2200) | 0 vs 9 | 1 | ---- |
| Palomba | 2011 | Italy | 1012 | 403 | 609 | ---- | 199 (150-310)  135 (105-160) | 134 vs 376 | 200 (50-850)  550 (150-1800) | ---- | 53 | 83 vs 154 |
| Malzoni | 2008 | Italy | 159 | 81 | 78 | 2.1 +/- 0.5  5.1 +/- 1.2 | 136 +/- 31  123 +/- 29 | 9 vs 11 | 50 +/- 12 (20-90)  145 +/- 35 (60-255) | 0 | 0 | 7 vs 9 |
| Malur | 2001 | Germany | 70 | 37 | 33 | 8.6 +/- 2.7  11.7 +/- 3.8 | 176.4 +/- 85.4  166.1 +/- 61 | 11 vs 13 | 229.2 +/- 190.2  594.2 +/- 629.9 | 1 vs 11 | 0 | 1 vs 2 |
| Leiserowitz | 2009 | USA | 12743 | 978 | 11765 | 2.40  4.36 | ---- | 334 vs 7869 | ---- | ---- | 154 | 6 vs 16 |
| Kalogiannidis | 2007 | Belgium | 169 | 69 | 100 | 5 (2-14)  8 (3-35) | 172 (60-360)  137 (60-330) | 5 vs 8 | 300 (100-1000)  355 (100-2100) | ---- | 4 | ---- |
| Howard | 1993 | USA | 30 | 15 | 15 | 3.7 +/- 1.7  5.2 +/- 3.1 | 69 +/- 36  119 +/- 25 | 2 vs 6 | 532 +/- 279  465 +/- 208 | ---- | ---- | ---- |
| Frigerio | 2006 | Italy | 110 | 55 | 55 | 4 vs. 8.5 | 220 (80-375)  175 (70-360) | 11 vs 16 | 285 (100-800)  177 (10-900) | ---- | 3 | ---- |
| Devaja | 2010 | UK | 182 | 74 | 108 | 4 (3-11)  6 (3-41) | 175 (85-205)  115 (75-255) | 3 vs 4 | ---- | ---- | 4 | ---- |
| Atabekoglu | 2004 | Turkey | 46 | 23 | 23 | 2.7 +/- 0.8  4.3 +/- 1.4 | 105.5 +/- 23.1  77.3 +/- 18.7 | ---- | 152 +/- 103.4  294.8 +/- 155.5 | ---- | ---- | ---- |
| Asgari | 2008 | Iran | 90 | 30 | 60 | 3.43 +/- 0.90  3.93 +/- 1.02 | 145.83 +/- 41.55  100.17 +/- 39.95 | 2 vs 16 | ---- | 1 vs 3 | ---- | ---- |
| Fram | 2002 | Australia | 61 | 29 | 32 | 2.3 vs. 5.5 | 136.2 vs. 101.9 | 3 vs 6 | 145.5 vs. 501.6 | 1 vs 2 | 2 | ---- |
| Falcone | 1998 | USA | 44 | 23 | 21 | 1.5 (1,2.3)  2.5 (1.5,2.5) | 180 (139-225)  130 (97-155) | 8 vs 5 | 450 (250-700)  250 (150-300) | 3 vs 4 | 1 | ---- |

* B. T = Blood Transfusion, H. S = Hospital Stay, C. = Complications (pre/intra/post-operative), B. L = Blood Loss (mL).

**Table 1**
